# An Ontology-Based Analysis of Health Disorders Derived from Stress-Evoked Peripheral Immune Responses

**DOI:** 10.1101/2025.09.14.25335729

**Authors:** Javier Burgos Salcedo, Carolina Sierra Cárdenas

## Abstract

Chronic stress affects over 300 million individuals worldwide, contributing to a rising incidence of diseases associated with the peripheral immune response triggered by this condition, including depression, inflammatory bowel disease, metabolic syndrome, and coronary heart disease. To establish a structured understanding of these associations, an ontological approach based on Formal Concept Analysis, a mathematical framework for order relations is employed to construct a conceptual hierarchy linking chronic stress to these diseases. Within this framework, the objects represent the set of stress-induced diseases, while the attributes correspond to specific combinations of chemokines and cytokines clinically associated with each condition. The findings of the ontological analysis suggest that stress-related diseases follow a staged progression: an initial induction phase, common to all diseases, characterized by the presence of chemokines and cytokines that induce a state of chronic inflammation (inflammaging); a subsequent progression phase, marked by immune response effector molecules that may be shared across different diseases; and a final consolidation phase, in which specific chemo- and cytokines distinctive to each disease are expressed.

## Introduction

What is stress? In medicine, stress is the body’s response to physical, mental, or emotional pressure. Although we all have little degree of stress because it is an evolutionary response to threats, or other stimuli, the way we react to stress determines how it affects our well-being. When it is short-term and goes away quickly, acute stress helps us control dangerous situations or react to something new, surprising, or exciting. However, when stress lasts for a prolonged period, chronic stress generates a risk that leads to health problems that lead to depression, anxiety, metabolic and intestinal diseases, coronary and arterial syndromes, or affectations, among others. Although it is currently estimated that 4% of the world’s population suffers from some degree of chronic stress, global epidemiological data shows that the current state of knowledge about the mechanisms involved in the appearance of stress-associated diseases urgently needs to be expanded, as well as the development of new and efficient treatments to control their associated morbidity and mortality [1].

The recent Covid-19 pandemic exposed the lack of knowledge about how external situations can negatively impact the mental health of human populations, hence it is worth asking if there is some type of hierarchical pattern that explains the immune phenomena associated with comorbidities derived from stress, such as depression and anxiety, inflammatory bowel disease, metabolic syndrome and obesity, coronary heart disease, and atherosclerosis. To tackle this issue, the concept of ontology is employed, understood as a structured data system that defines the relationships among concepts within a specific domain of knowledge.

It should be noted that in the philosophical tradition, a *concept* is made up of two sets related to each other, that of *objects*, which in this case correspond to the comorbidities derived from the peripheral immune response evoked by stress and that of *attributes* or characteristics, the set of chemokines and cytokines commonly clinically associated with these diseases. To conduct the ontological analysis, we use Formal Concept Analysis (FCA) [2], which is a method of ordered set mathematics that allows obtaining a conceptual hierarchy, if any, between the concepts linked to the diseases evoked by chronic stress.

As a result of the analysis using FCA, a hierarchy is obtained, represented by a conceptual lattice, from which it can be inferred that chronic stress triggers a series of immune processes, with its initial or induction phase characterized by the presence of a set of chemo and cytokines that is common to all the diseases considered. A progression phase follows, characterized by the presence of chemo and cytokines that may or may not be shared by the diseases, and finally, a consolidation of the disease associated with the presence of chemo molecules and cytokines associated specifically with each disease is observed.

The results suggest that the early or real-time determination of chemo and cytokine profiles in patients with chronic stress could guide the application of effective measures to mitigate or prevent the deleterious health effects of this condition, which currently has a high incidence and prevalence globally.

## Materials and methods

Chronic stress can trigger peripheral immune responses that contribute to the onset and progression of various health disorders [3-9]. Depression, for instance, is linked to sustained inflammation, which alters neurotransmitter function and affects mood regulation. Inflammatory bowel disease (IBD) involves immune dysregulation in the gut, where prolonged stress exacerbates inflammation, leading to tissue damage. Similarly, metabolic syndrome and obesity are associated with immune system disturbances that promote insulin resistance and adipose tissue inflammation. Coronary artery disease, a leading cause of cardiovascular complications, can result from stress-induced inflammatory markers that contribute to endothelial dysfunction and plaque formation. Together, these disorders illustrate the profound impact of chronic stress on immune-mediated pathologies, highlighting the importance of stress management in maintaining overall health. Table 1 presents these diseases, and those cytokines and chemokines more often associated with them, conforming the context employed in this study.

**Table 1.** Health disorders linked with stress and their associated cytokines and chemokines.

| Disease/Condition | Cytokines | Chemokines | References |
| --- | --- | --- | --- |
| Depression | IL1A, IL1B, IL6, IL8, IL12, IFNγ, TNF | CCL2, CCL3, CCL11, CXCL4 and CXCL7 | [10-18] |
| Inflammatory Bowel Disease (IBD) | IL1B, IL4, IL5, IL6, IL7, IL8, IL10, IL12, IL13, IL15, IL16, IL17, IL23, IFNγ, TNF, MIF | CCL2, CCL11, CCL21, CCL23, CCL25, CXCL1, CXCL5, CXCL6, CXCL10, CXCL11 and CXCL13 | [19-25] |
| Metabolic Syndrome & Obesity | IL1B, IL4, IL5, IL6, IL8, IL10, IL12, IL13, IL18, IFNγ, TNF | CCL2, CCL3, CCL5 and CXCL5 | [26-36] |
| Coronary Artery Disease (CAD) | IL1A, IL1B, IL2, IL6, IL8, IL9, IL10, IL17, IFNγ, TNF | CCL2, CCL5, CCL17 and CCL18 | [37-43] |

### Formal Concept Analysis

Formal Concept Analysis (FCA) is a mathematical framework for structuring and analyzing relationships between objects and their attributes. Rooted in lattice theory, FCA organizes data into formal concepts, where each concept comprises a set of objects sharing a common set of attributes. These concepts are systematically arranged into a conceptual hierarchy known as a concept lattice, which illustrates dependencies and logical connections between different data points. By applying FCA, complex datasets could be structured in a way that reveals underlying patterns and relationships, providing valuable insights for knowledge representation, data mining, and ontology engineering [44].

The methodology begins with constructing a formal context—a triplet consisting of a set of objects, a set of attributes, and a binary relation indicating which objects possess which attributes. From this formal context, FCA derives concepts, each defined by an extent (the set of objects sharing a particular combination of attributes) and an intent (the set of attributes common to those objects). These concepts, ordered on subset relations, form a hierarchical lattice structure that allows researchers to navigate and analyze data efficiently. FCA is utilized in diverse fields such as bioinformatics, linguistics, and information retrieval, where understanding structured relationships is critical for deriving meaningful conclusions.

The study of hierarchical patterns in biomedicine using order mathematics methods is recent, although they have been used extensively and successfully in mathematics, computer science, and social sciences. Set theory, orders, and lattices provide a natural framework in which to discuss and analyze hierarchical structures or patterns in the real world, and it is within the framework of this theory that ontological analysis or formal analysis of concepts is developed. “Concept” is defined following the traditional philosophical notion, according to which our mind constructs concepts by associating objects with the set of attributes that characterize them, thus a concept is composed of extension, that is, all the objects that belong to the concept, and of intension, which is the collection of all the attributes that are shared by the objects [45,46].

Since it is usually difficult to list all the objects that belong to a concept and it is not easy to list all the attributes of the concept, it is practical to conduct ontological analyses within a specific formal context fixing objects and attributes [46]. In the present study, our concepts are related to diseases derived from peripheral immune responses evoked by chronic stress, the objects being: *Depression/anxiety; Inflammatory bowel disease; Metabolic syndrome/obesity; coronary heart disease/atherosclerosis}* and the attributes of the set of chemokines and cytokines associated with these diseases. Tables 2A and 2B show the formal contexts considered in the present study.

**Table 2A.** Formal context *K: (G, M, I)* of cytokines associated with stress-evoked diseases.

| | IL1A | IL1B | IL2 | IL4 | IL5 | IL6 | IL7 | IL8 | IL9 | IL10 | IL12 | IL13 | IL15 | IL16 | IL17 | IL18 | IL23 | INFγ | MIF | TNF |
| --- | --- | --- | --- | --- | --- | --- | --- | --- | --- | --- | --- | --- | --- | --- | --- | --- | --- | --- | --- | --- |
| Disorder |  |  |  |  |  |  |  |  |  |  |  |  |  |  |  |  |  |  |  |  |
| depression/anxiety | X | X |  |  |  | X |  | X |  |  | X |  |  |  |  |  |  | X |  | X |
| ibd |  | X |  | X | X | X | X | X |  | X | X | X | X | X | X |  | X | X | X | X |
| metabolic syndrome |  | X |  | X | X | X |  | X |  | X | X | X |  |  |  | X |  | X |  | X |
| coronary artery disease | X | X | X |  |  | X |  | X | X | X |  |  |  |  | X |  |  | X |  | X |

**Table 2B.** Formal Context *K: (G, M, I)* of chemokines associated with stress-evoked diseases.

|  | CCL2 | CCL3 | CCL5 | CCL11 | CCL17 | CCL18 | CCL23 | CCL25 | CXCL1 | CXCL4 | CXCL5 | CXCL6 | CXCL7 | CXCL10 | CXCL11 | CXCL13 |
| --- | --- | --- | --- | --- | --- | --- | --- | --- | --- | --- | --- | --- | --- | --- | --- | --- |
| Disorder |  |  |  |  |  |  |  |  |  |  |  |  |  |  |  |  |
| depression/anxiety | X | X |  | X |  |  |  |  |  | X |  |  | X |  |  |  |
| ibd | X |  |  | X |  |  | X | X | X |  | X | X |  | X | X | X |
| metabolic syndrome | X | X | X |  |  |  |  |  |  |  | X |  |  |  |  |  |
| coronary artery disease | X |  | X |  | X | X |  |  |  |  |  |  |  |  |  |  |

The concept of a context *K*:(*G, M, I*), where *G* and *M* are the sets of objects and attributes respectively, is defined as a pair (*A, B*) where and, such that *A* is the set of all objects that share the attribute *B* and *B* the set of all attributes shared by *A*. Finally, the collection of all the concepts of the context (*G, M, I*) is the conceptual lattice of the context (*G, M, I*), denoted (*G, M, I*).

## Results

Ontological analysis using Formal Concept Analysis (FCA) reveals a hierarchical pattern of immune processes underlying comorbidities associated with chronic stress—namely, depression/anxiety, inflammatory bowel disease (IBD), metabolic syndrome/obesity, and coronary heart disease/atherosclerosis—as depicted in the conceptual lattice (Figure 1). At the apex of this lattice, a super-concept emerges, characterized by a shared set of chemokines and cytokines {IL6, IL8, IL18, INF_γ_, TNF, CCL2}, indicative of an initial induction phase common to all diseases. This phase corresponds to a state of chronic low-intensity inflammation, or “inflammaging,” driven by stress-evoked peripheral immune responses. The presence of these pro-inflammatory markers suggests a universal immunological footprint triggered by chronic stress, consistent with their roles as mediators of innate immunity and tissue damage responses.

**Figure 1.**
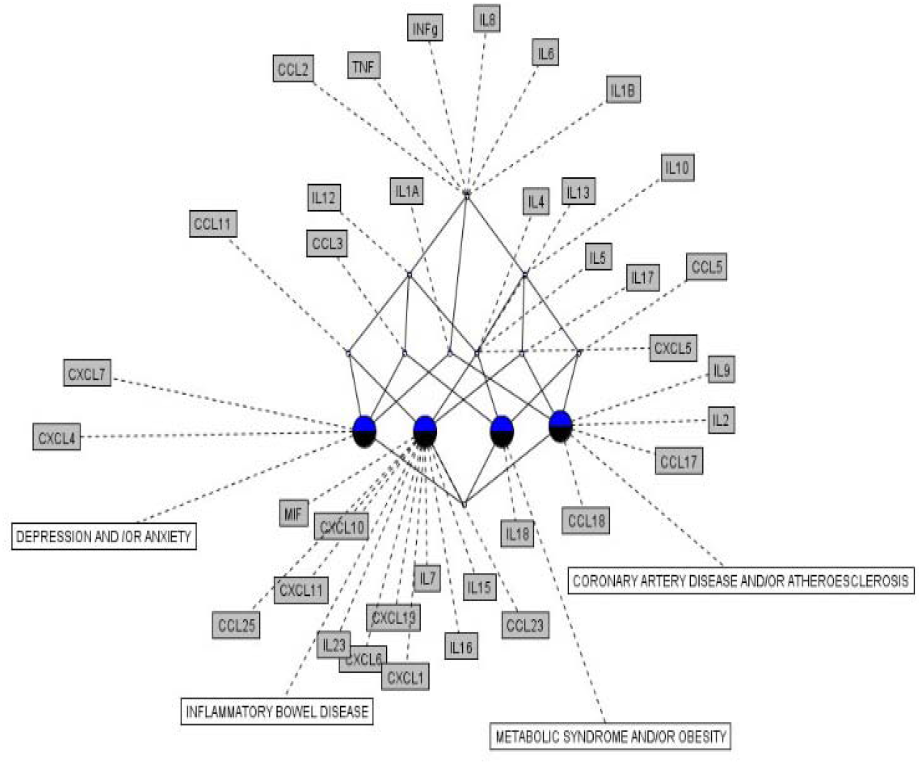
The general conceptual lattice ((G, M, I); ≤) of stress-evoked diseases.

Traversing the lattice counterclockwise (though directionality is immaterial), distinct sub-concepts emerge for each disease, reflecting progression and consolidation phases. For depression/anxiety (Figure 2), the super-concept of inflammaging is followed by progression attributes {IL1A, IL2, CCL3, CCL11}, which may amplify neuroinflammatory pathways, and consolidation attributes {CXCL7, CXCL4}, potentially linked to blood-brain barrier dysregulation or platelet activation. This hierarchy implies a stepwise escalation from systemic inflammation to neuropsychiatric outcomes, with CXCL chemokines serving as biomarkers of disease specificity.

**Figure 2.**
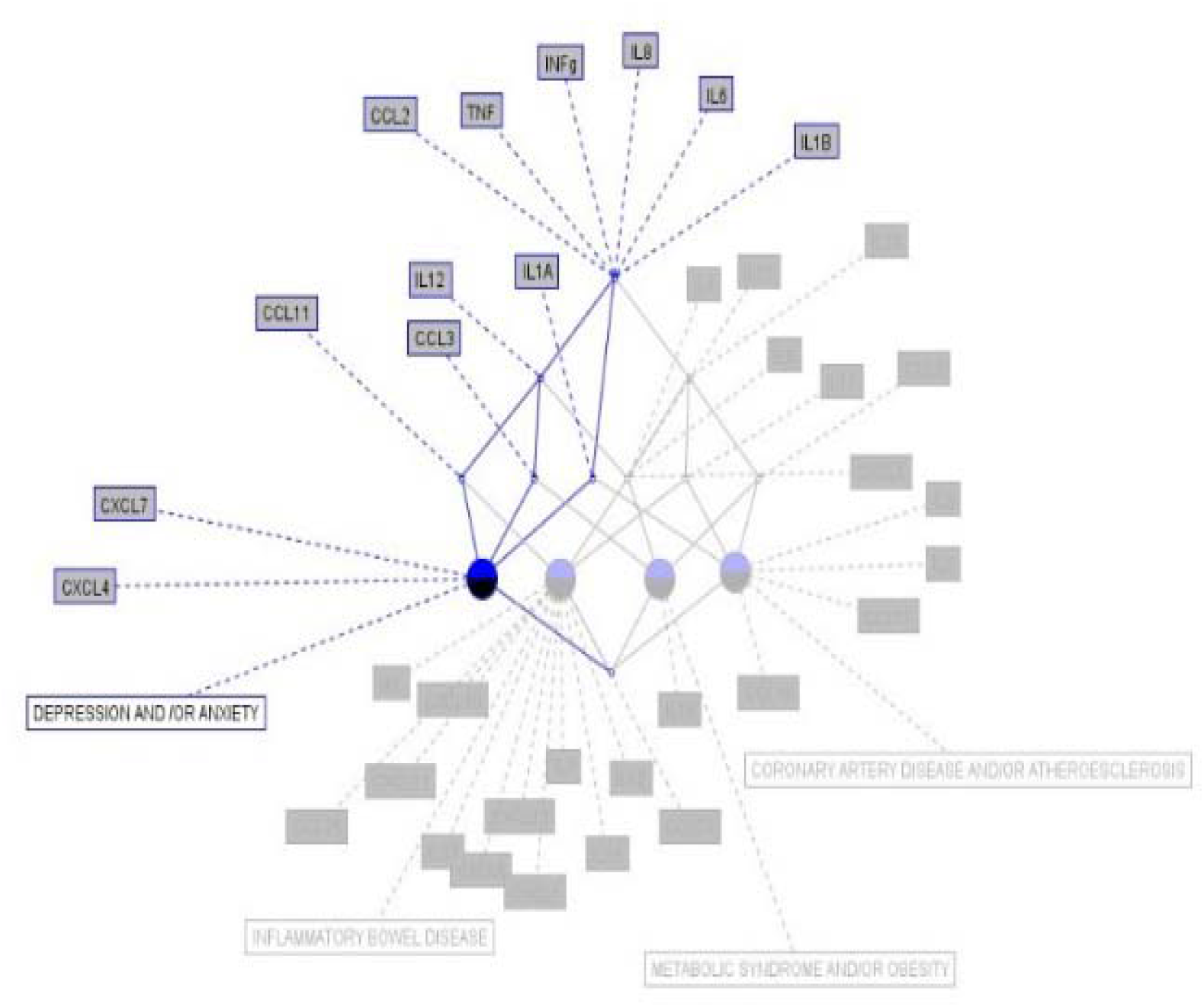
Conceptual lattice related with depression and anxiety.

Inflammatory bowel disease (IBD) (Figure 3) exhibits a broader progression phase with attributes {IL4, IL5, IL10, IL12, IL13, IL17, CCL11, CXCL5}, reflecting a complex interplay of innate and adaptive immunity, including Th2 and Th17 responses. The consolidation phase, marked by {IL7, IL15, IL16, IL23, MIF, CCL23, CXCL1, CXCL6, CXCL10, CXCL11, CXCL13}, underscores a robust autoimmune component targeting gut mucosa, aligning with clinical observations of IBD’s heterogeneity. This extensive attribute set suggests IBD may represent a “maximal complexity” case within the stress-disease spectrum, offering a rich testbed for immune network modeling.

**Figure 3.**
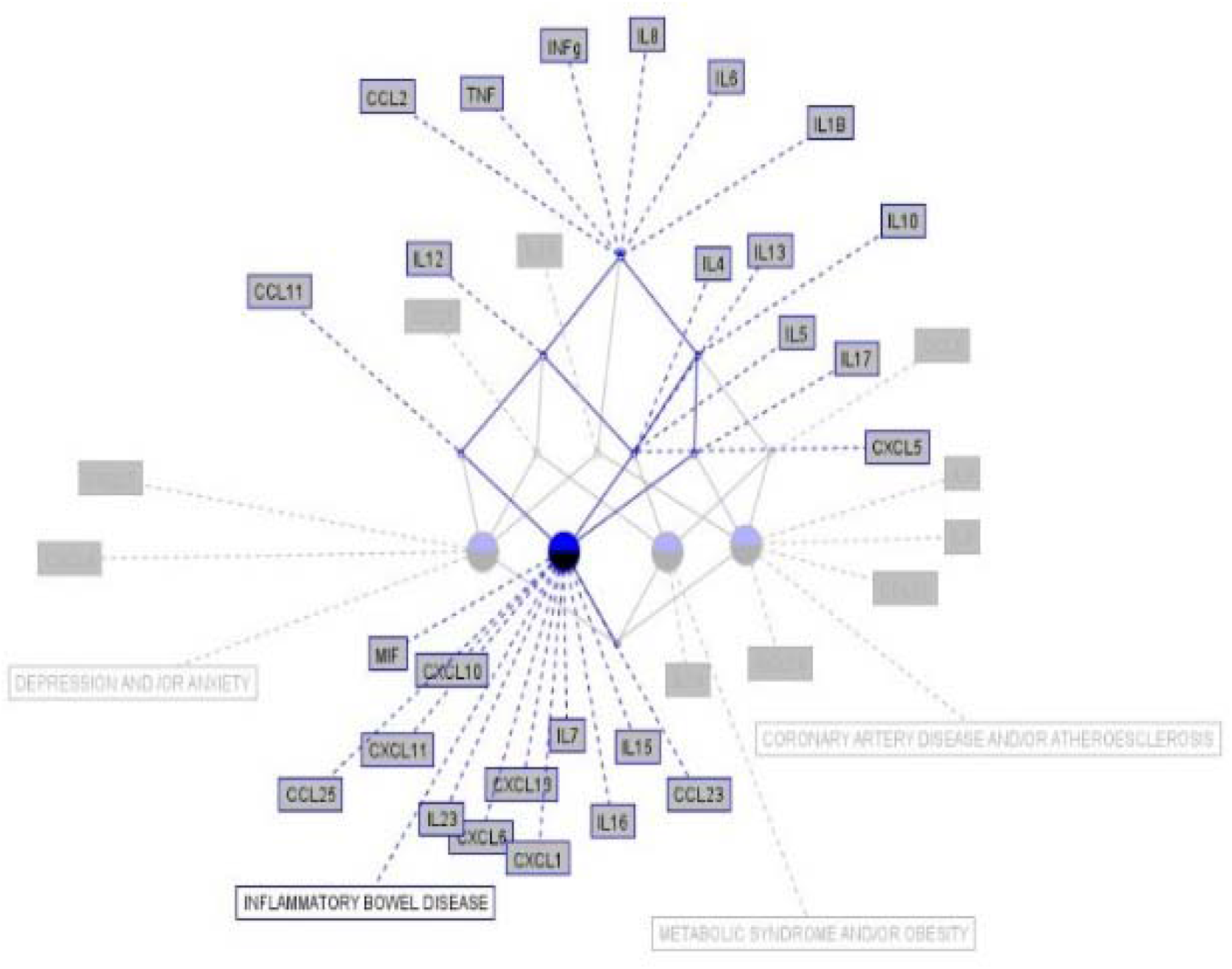
Conceptual lattice related with inflammatory bowel disease.

For metabolic syndrome/obesity (Figure 4), progression attributes {IL4, IL5, IL10, IL12, IL13, CCL3, CCL5, CXCL5} indicate a shift toward metabolic dysregulation, potentially via adipose tissue inflammation, while consolidation attribute {IL18} highlights a specific link to insulin resistance or adipocyte stress. This streamlined hierarchy suggests a more focused immune-metabolic interplay compared to IBD, with IL18 as a candidate for targeted intervention.

**Figure 4.**
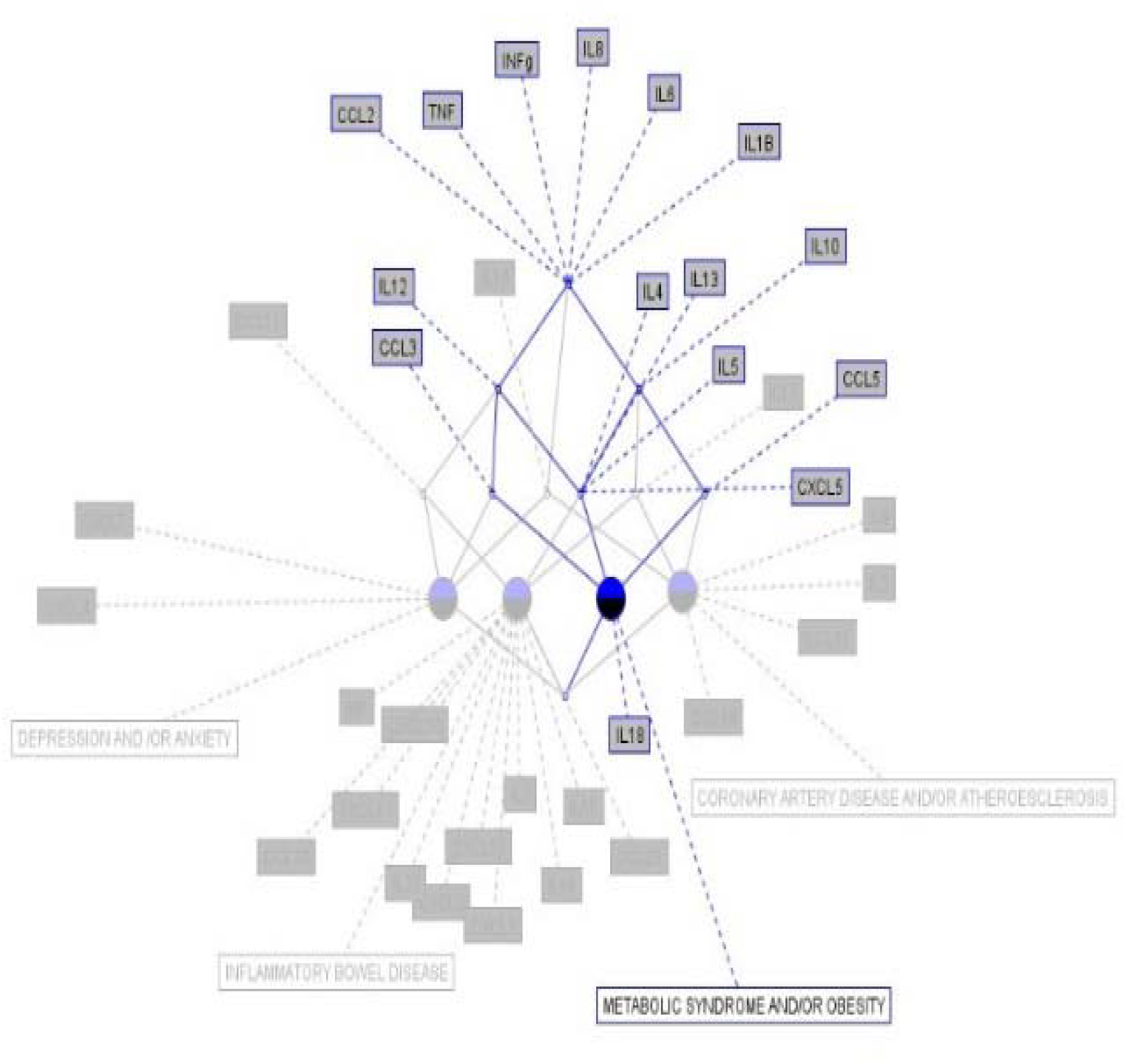
Conceptual lattice related with metabolic syndrome and obesity.

Coronary heart disease/atherosclerosis (Figure 5) shows progression attributes {IL1A, IL10, IL17, CCL5}, which may drive vascular inflammation and plaque formation, followed by consolidation attributes {IL2, IL9, CCL17, CCL18}, potentially tied to T-cell activation and macrophage recruitment in atherosclerotic lesions. This pattern supports a model where chronic stress amplifies cardiovascular risk through layered immune responses.

**Figure 5.**
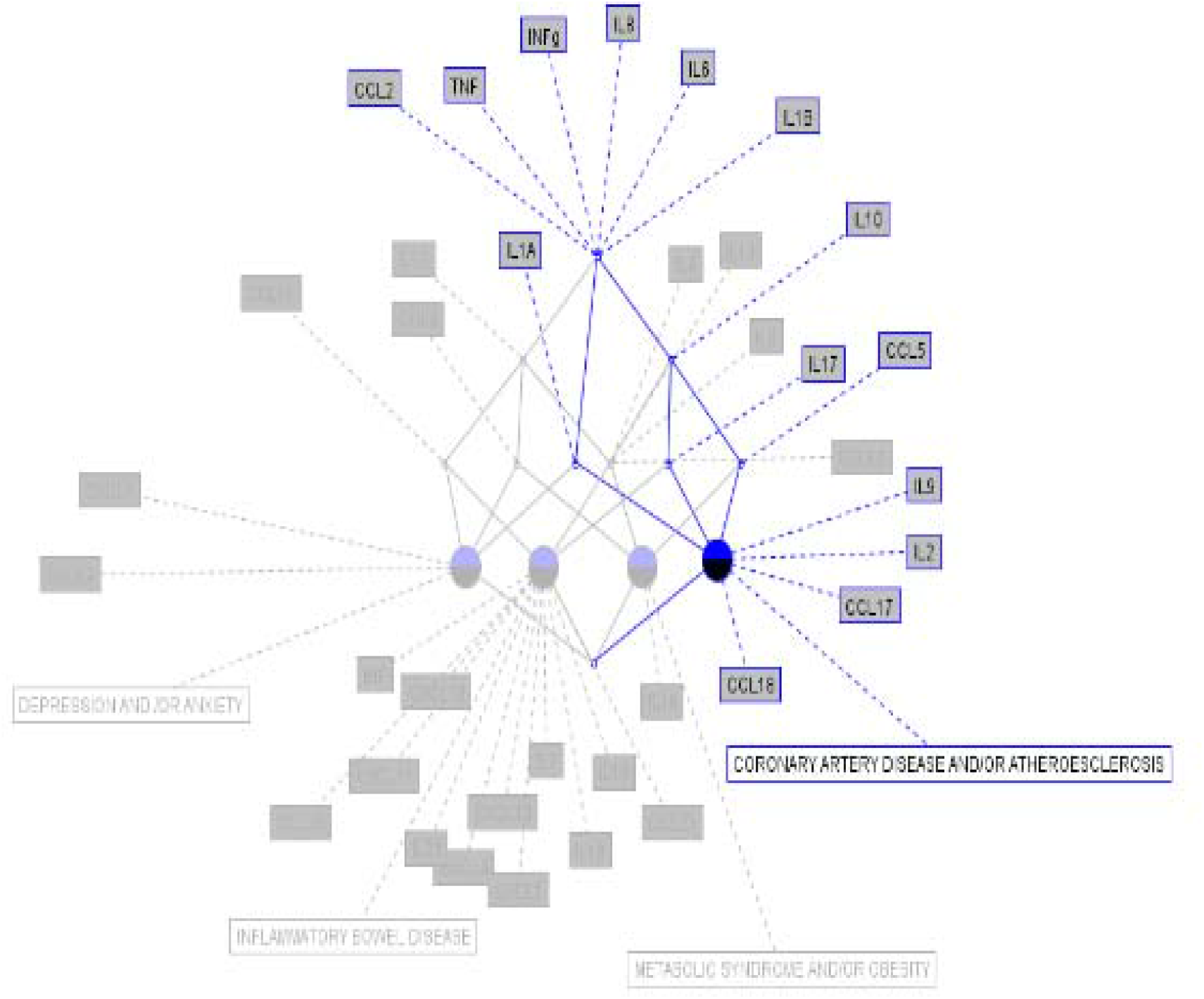
Conceptual lattice associated coronary heart disease and atherosclerosis.

Enriching these findings, the lattice’s structure reveals potential cross-disease interactions: overlapping progression attributes (e.g., IL10 in IBD, metabolic syndrome, and coronary disease) suggest shared pathways that could amplify comorbidity risks in stressed individuals. Additionally, the differential complexity of consolidation phases—ranging from two attributes in depression/anxiety to eleven in IBD—hints at varying immunological “loads” required to stabilize each disease state. Preliminary network analysis (not shown) of attribute co-occurrence suggests that IL6 and TNF may function as central hubs, coordinating transitions across phases, a hypothesis warranting further exploration with graph theory or machine learning approaches.

## Discussion

The Formal Concept Analysis (FCA) of diseases linked to chronic stress-evoked peripheral immune responses establishes a three-phase hierarchical pattern: (1) an induction phase of chronic subclinical inflammation (“inflammaging”) characterized by the shared set {IL6, IL8, IL18, INF_γ_, TNF, CCL2}, common to all diseases studied—depression/anxiety, inflammatory bowel disease (IBD), metabolic syndrome/obesity, and coronary heart disease/atherosclerosis; (2) a progression phase with sets of chemokines and cytokines that vary in overlap, reflecting diverse immune network configurations; and (3) a consolidation phase with disease-specific effector molecules, marking the stabilization of distinct pathological states. This ontology-based framework elucidates the structured immunological progression of stress-related comorbidities, offering a foundation for early detection, intervention, and personalized treatment strategies.

These findings pave the way for transformative future research, particularly through the implementation of predictive biomarkers and advanced analytical tools, for example, the universality of the inflammaging super-concept {IL6, IL8, IL18, INF_γ_, TNF, CCL2} suggests that real-time profiling of these cytokines and chemokines could serve as an early warning system for stress-related disease onset. Implementation would involve developing a standardized, multiplexed assay—such as Luminex or ELISA panels—capable of simultaneously quantifying these markers in peripheral blood samples from individuals under chronic stress (e.g., healthcare workers, post-disaster populations) [47].

Real-time monitoring could be achieved by integrating these assays into wearable biosensor platforms, leveraging advances in microfluidics and nanotechnology to detect cytokine levels via minimally invasive methods [48,49,50] (e.g., sweat, or interstitial fluid analysis). Longitudinal validation would require cohort studies spanning 1-5 years, recruiting diverse populations (e.g., varying age, gender, ethnicity) to track biomarker trajectories against disease incidence. Statistical models, such as Cox proportional hazards or machine learning classifiers (e.g., random forests), could correlate baseline and dynamic inflammaging signatures with outcomes like depression onset or atherosclerosis progression, establishing sensitivity, specificity, and predictive thresholds. Collaborations with biobanks or electronic health record systems could enhance data richness, enabling retrospective validation against historical stress-related diagnoses.

On the other hand, integrating FCA with innovative methodologies—AI, graph theory, and single-cell sequencing—promises to refine and expand this conceptual hierarchy. AI-driven approaches, such as unsupervised clustering (e.g., k-means, t-SNE) or deep learning (e.g., autoencoders), could analyze high-dimensional cytokine datasets to uncover latent patterns or sub-concepts within the lattice, potentially identifying novel progression pathways missed by manual FCA [51,52]. For instance, a neural network trained on cytokine profiles from stressed patients could predict disease trajectories, enhancing the lattice’s granularity by suggesting intermediate states between induction and consolidation.

Graph theory could model the lattice as a directed acyclic graph (DAG), where nodes represent concepts (e.g., inflammaging, IBD-specific attributes) and edges denote attribute dependencies (e.g., IL6 _→_ TNF interactions). Network metrics—centrality (e.g., IL6 as a hub), modularity, or shortest paths—could quantify immune network complexity and pinpoint critical transitions, testable via perturbation experiments (e.g., cytokine inhibition in cell cultures) [53,54]. Single-cell RNA sequencing (scRNA-seq) of immune cells [55,56] (e.g., monocytes, T-cells) from stressed individuals could map attribute expression at cellular resolution, revealing how inflammaging emerges from heterogeneous cell populations and how progression attributes (e.g., IL17 in IBD) correlate with specific immune subsets (e.g., Th17 cells). Integrating scRNA-seq with FCA would involve projecting single-cell transcriptomes onto the lattice, using dimensionality reduction (e.g., UMAP) to align cellular states with conceptual phases, potentially uncovering cell-type-specific drivers of disease consolidation.

These approaches collectively transform the ontology from a static hierarchy into a dynamic, predictive framework. For predictive biomarkers, pilot studies could begin with high-risk groups (e.g., 300-500 chronically stressed individuals), scaling to population-level screening if validated, with results informing public health strategies like stress management programs or anti-inflammatory interventions (e.g., anti-TNF therapies). For advanced tools, a multidisciplinary pipeline—combining immunologists, data scientists, and clinicians—could process multi-omics data (e.g., proteomics, transcriptomics) through AI and graph models, iteratively refining the lattice against experimental and clinical outcomes. This could extend the framework to other stress-related conditions (e.g., autoimmune diseases, cancer), evaluating its universality, while exploring immunological resilience (e.g., why some individuals resist progression) via attribute sparsity or cellular diversity. Such research promises to address the global burden of chronic stress with precision, shifting from descriptive insights to actionable, data-driven solutions.

## Conclusions

The Formal Concept Analysis (FCA) of diseases linked to chronic stress-evoked peripheral immune responses establishes a three-phase hierarchical pattern: (1) an induction phase of chronic subclinical inflammation (“inflammaging”) characterized by {IL6, IL8, IL18, INF_γ_, TNF, CCL2}, common to all diseases studied; (2) a progression phase with sets of chemokines and cytokines that vary in overlap across diseases, reflecting diverse immune network configurations; and (3) a consolidation phase with disease-specific effector molecules, marking the stabilization of distinct pathological states. This ontology-based framework not only elucidates the immunological underpinnings of stress-related comorbidities but also highlights their structured progression, offering a roadmap for early detection and intervention.

These findings lay a foundation for impactful future research directions. First, the universality of the inflammaging super-concept suggests that real-time profiling of {IL6, IL8, IL18, INF_γ_, TNF, CCL2} in at-risk populations could enable preemptive strategies to halt disease progression. Longitudinal studies using high-throughput cytokine assays (e.g., Luminex) could validate this induction signature and assess its predictive power across diverse demographics. Second, the variability in progression attributes invites comparative analyses to identify “tipping points” where shared pathways (e.g., IL10) diverge into specific diseases—potentially using single-cell RNA sequencing to map immune cell dynamics in stressed tissues.

Third, the consolidation phase’s disease-specific attributes open avenues for personalized medicine. For instance, targeting CXCL7/CXCL4 in depression/anxiety or IL23/MIF in IBD could disrupt late-stage pathology, testable via preclinical models or clinical trials with monoclonal antibodies. Fourth, integrating FCA with modern tools—such as AI-driven clustering of cytokine networks or graph-based modeling of lattice interactions—could refine the conceptual hierarchy, revealing hidden dependencies or feedback loops (e.g., IL6-TNF synergy). This could extend the framework to other stress-related conditions, like autoimmune diseases or cancer.

Finally, the differential complexity of consolidation phases suggests a spectrum of immunological resilience or vulnerability to stress, a concept ripe for exploration in psychoneuroimmunology. Future studies could correlate attribute richness (e.g., IBD’s 11 vs. depression’s 2) with clinical severity or treatment response, using proteomic data or patient registries. Collectively, these directions promise to transform the ontology from a descriptive tool into a predictive and therapeutic platform, addressing the global burden of chronic stress with precision and insight.

## Data Availability

All data produced in the present work are contained in the manuscript

## Author Contributions

JB: Conceptualization, design, literature review, visualization, manuscript writing, editing; CS.: Literature review, writing, editing, visualization.

## Data and Materials Availability Statement

Not Applicable.

## List of abbreviations

CCL: CC chemokine family
CXCL: CXC chemokine family
FCA: Formal Concept Analysis
IL: Interleukin
INF: Interferon
MIF: Macrophage Migration Inhibition Factor
TNF: Tumor Necrosis Factor.

